# Comparative sensitivity of Early Cystic Fibrosis Lung Disease detection tools in school aged children

**DOI:** 10.1101/2023.11.05.23298077

**Authors:** Katie J Bayfield, Oliver Weinheimer, Anna Middleton, Christie Boyton, Rachel Fitzpatrick, Brendan Kennedy, Anneliese Blaxland, Geshani Jayasuriya, Neil Caplain, Mark O Wielpütz, Lifeng Yu, Craig J Galban, Terry E Robinson, Brian Bartholmai, Per Gustafsson, Dominic Fitzgerald, Hiran Selvadurai, Paul D Robinson

## Abstract

**Background:** Effective detection of early lung disease in cystic fibrosis (CF) is critical to understanding early pathogenesis and evaluating early intervention strategies. We aimed to compare ability of several proposed sensitive functional tools to detect early CF lung disease as defined by CT structural disease in school aged children.

**Methods:** 50 CF subjects (mean±SD 11.2±3.5y, range 5-18y) with early lung disease (FEV_1_≥70% predicted: 95.7±11.8%) performed spirometry, Multiple breath washout (MBW, including trapped gas assessment), oscillometry, cardiopulmonary exercise testing (CPET) and simultaneous spirometer-directed low-dose CT imaging. CT data were analysed using well-evaluated fully quantitative software for bronchiectasis and air trapping (AT).

**Results:** CT bronchiectasis and AT occurred in 24% and 58% of patients, respectively. Of the functional tools, MBW detected the highest rates of abnormality: S_cond_ 82%, MBW_TG RV_ 78%, LCI 74%, MBW_TG IC_ 68% and S_acin_ 51%. CPET VO_2_peak detected slightly higher rates of abnormality (9%) than spirometry (FEV_1_/FVC 8%, FEF_25-75_ 8%, FEV_1_ 2%). For oscillometry AX (14%) performed better than Rrs (2%) whereas Xrs and R5-19 failed to detect any abnormality. LCI and S_cond_ correlated with bronchiectasis (r=0.55-0.64, p<0.001) and AT (r=0.73-0.74, p<0.001). MBW-assessed trapped gas was detectable in 92% of subjects and concordant with CT-assessed AT in 74%.

**Conclusions:** Significant structural and functional deficits occur in early CF lung disease, as detected by CT and MBW. For MBW, additional utility, beyond that offered by LCI, was suggested for S_cond_ and MBW-assessed gas trapping. Our study reinforces the complementary nature of these tools and the limited utility of conventional oscillometry and CPET in this setting.

**AUTHOR CONTRIBUTIONS:**

- Conception and design: KB, OW, MOW, BB, TER, PG, DF, HS, PR
- Acquisition, analysis and interpretation of data: KB, OW, CB, AB, RF, PG, AM, HS, BK, MOW, LY, CG, BB, TER, PR
- Writing the manuscript or revising it critically for important intellectual content: KB, OW, CB, AB, RF, AM, BK, GJ, NC, PG, DF, HS, CG, MOW, BB, TER, PR

**HIGHLIGHTS:**

- In early lung disease, multiple breath washout and CT are complementary tools
- MBW and CT detect more abnormality than oscillometry, CPET and spirometry
- Sensitivity of MBW to detect abnormality can be increased by S_cond_ and MBW-assessed trapped gas

**GRANT SUPPORT:** Australian Cystic Fibrosis Research Trust: 2018 Innovation Grant. This study was supported by grants from the German Federal Ministry of Education and Research (82DZL004A1) (to OW, MOW).

## INTRODUCTION

Cystic fibrosis (CF) lung disease occurs early in life(1) and is progressive(2). Insensitivity of FEV_1_ to detect early disease is well recognised in people with CF (pwCF), with median FEV_1_%predicted within the normal range during childhood. With CFTR modulator therapy now available for ≈90% of pwCF, the proportion with FEV_1_ values within the normal range will increase across both paediatric and adult pwCF(3). Despite normal FEV_1_, computed tomography (CT)(4) detects significant structural abnormalities, and early structural disease predicts later disease outcomes(5), including lung function trajectory(6, 7). Optimal characterisation of early structural and functional abnormalities, and their relationships, has been highlighted as critical to future efforts to detect and prevent disease progression(8). Detailed studies focused on early disease are therefore critical to advance current understanding of how best to use more sensitive functional and imaging tools to monitor early disease, and ensure any increase in monitoring does not negatively impact existing care burden(9).

Current gold standard for early structural lung disease identification is CT(6, 10), performed under spirometer-direction to standardise lung volume acquisition(11-13). Automated fully quantitative assessment of airway changes has started to be implemented(14-19). To address concerns regarding cumulative effective radiation dose exposure in pwCF with increasing life expectancy (20) understanding the comparative utility of other functional tools is critical to define optimal surveillance approaches(8). Multiple breath washout (MBW) derived lung clearance index (LCI) has been shown to correlate with CT structural disease across a wide range of disease severity, based on FEV_1_ (21, 22). Modifications to MBW testing to generate phase III slope derived indices (S_cond_ and S_acin_)(23) and to detect trapped gas (TG) have been described(24) but rarely incorporated in studies focused on early structural lung disease alone. In other settings, oscillometry and cardio-pulmonary exercise testing (CPET) also offer improved sensitivity to detect disease, compared to spirometry, but how these tools all compare with MBW and CT remains unclear.

We therefore performed a direct comparison of several tools (MBW, oscillometry, CPET) to spirometry in the specific setting of early CF lung disease in a school-aged cohort of CF, with a gold standard definition of structural lung disease based on fully quantitative CT analysis. We hypothesized that in a direct comparison, novel approaches to MBW data analysis would provide the strongest ability to detect structural disease in early CF lung disease.

## METHODS

### Study Design and Participants

Inclusion criteria for this prospective cross-sectional study were: i) confirmed CF diagnosis from CFTR genotype(25), ii) age ≥5years, iii) FEV_1_ ≥70%predicted, iv) exacerbation free(26), and v) indication for routine surveillance CT by local standards. Ethics committee approval from the Sydney Children’s Hospitals Network Human Research Ethics Committee (SCHN HREC, ethics approval 18/SCHN/469) and written informed consent/assent was obtained from all parents/guardians and children. Further methodology is provided in the online supplement (OLS). A separate analysis comparing structural disease detected by conventional low dose and ultra-low dose CT within this cohort has recently been published elsewhere(27).

### Low dose computed tomography

Paired inspiratory-expiratory chest CT scans were performed on a third-generation dual-source scanner (SOMATOM Force, Siemens Healthineers AG, Germany) using a conventional low dose scanning protocol (see OLS). Spirometry-direction targeted published acceptability criteria(13) across consecutive inspiratory and expiratory scans (Figure E1 OLS). Scans were reconstructed at Kernel Br49d\3, slice thickness 0.6mm and 0.3mm increment. Preliminarily validated automatic software (YACTA v2.9.1.12) fully analysed all CT data to generate a range of structural lung disease variables (see OLS) including air trapping (AT variable A_1_, A_2_ and A_3_)(28, 29), airway wall thickness (WT) and bronchiectasis (Bronchiectasis index, BEI(30)).

### Lung function and cardio-pulmonary assessment

MBW and spirometry were performed in all subjects, oscillometry in those without tier III infections (due to inability to fully sterilise equipment) and CPET in those ≥8years. Nitrogen (N_2_) based MBW (Exhalyzer D, ECO MEDICS AG, Switzerland) was performed in triplicate(31), reporting LCI, S_cond_, S_acin_, visible trapped gas during a series of five inspiratory capacity (IC) breaths and after an additional vital capacity breath to residual volume (RV) (MBW_TG IC_; MBW_TG RV_). Oscillometry was performed as triplicate 60s recordings at 5-37Hz (tremoFlo® C-100 Airwave Oscillometry System™, THORASYS®, Canada)(32), reporting mean respiratory system resistance (Rrs) and reactance (Xrs) at 5Hz, Area under reactance curve (AX) and frequency dependence of resistance (Rrs5-19Hz). Spirometry was performed using JAEGER® (Vyntus Masterscreen PNEUMO, Germany) according to ATS/ERS recommendations(33, 34). CPET was performed using a Trackmaster® TMX428CP treadmill, Vyntus® CPX metabolic cart and the Bruce protocol(35), reporting oxygen uptake (VO_2_), at Anaerobic Threshold and Peak exercise (VO_2AT_; VO_2peak_).

### Statistical analysis

Data were analyzed using GraphPad Prism (version 8.4.3, GraphPad Software, USA) and R (version 4.0.2n, R Foundation for Statistical Computing, Austria). Abnormality for each index was defined as a z score of ±1.96 (e.g., spirometry and oscillometry) or used fixed values of upper limit of normal (e.g., CT variables, MBW, see OLS). Parametric data are presented as mean (SD) and nonparametric as median (range). Categorical data were compared with chi-square or Fisher’s exact tests, and continuous data compared with paired Student’s t-test or Wilcoxon signed-rank test. Structure-function relationships were explored using correlation (Pearson or Spearman r)(36), receiver operator characteristic (ROC) and multiple linear/logistic regression analysis. *p*<0.05 with correction for multiple comparisons, where appropriate, was considered statistically significant.

## RESULTS

### Study population

From 55 subjects, 50 were included in the final analysis: 5 were excluded due to i) CT export error (n=4), and ii) incorrect spirometer-directed technique (n=1). All children provided technically acceptable spirometry and MBW data (n=50), 86% for Oscillometry (n=48/50 eligible, 43/48 completed), and 63% for CPET (n=35/50 eligible, 23/35 completed). Demographics and characteristics of underlying CF at baseline are summarised in Table 1. The distribution of CFTR variants was 30% pF508.del homozygous, 66% pF508.del heterozygous and 4% minimal function without the pF508.del variant. Nutritional parameters were within the normal range with 76% of subjects pancreatic insufficient. Almost all were prescribed dornase-alfa and hypertonic saline, half were on CFTR modulator therapy, and approximately a third were on long-term azithromycin.

**Table 1.**
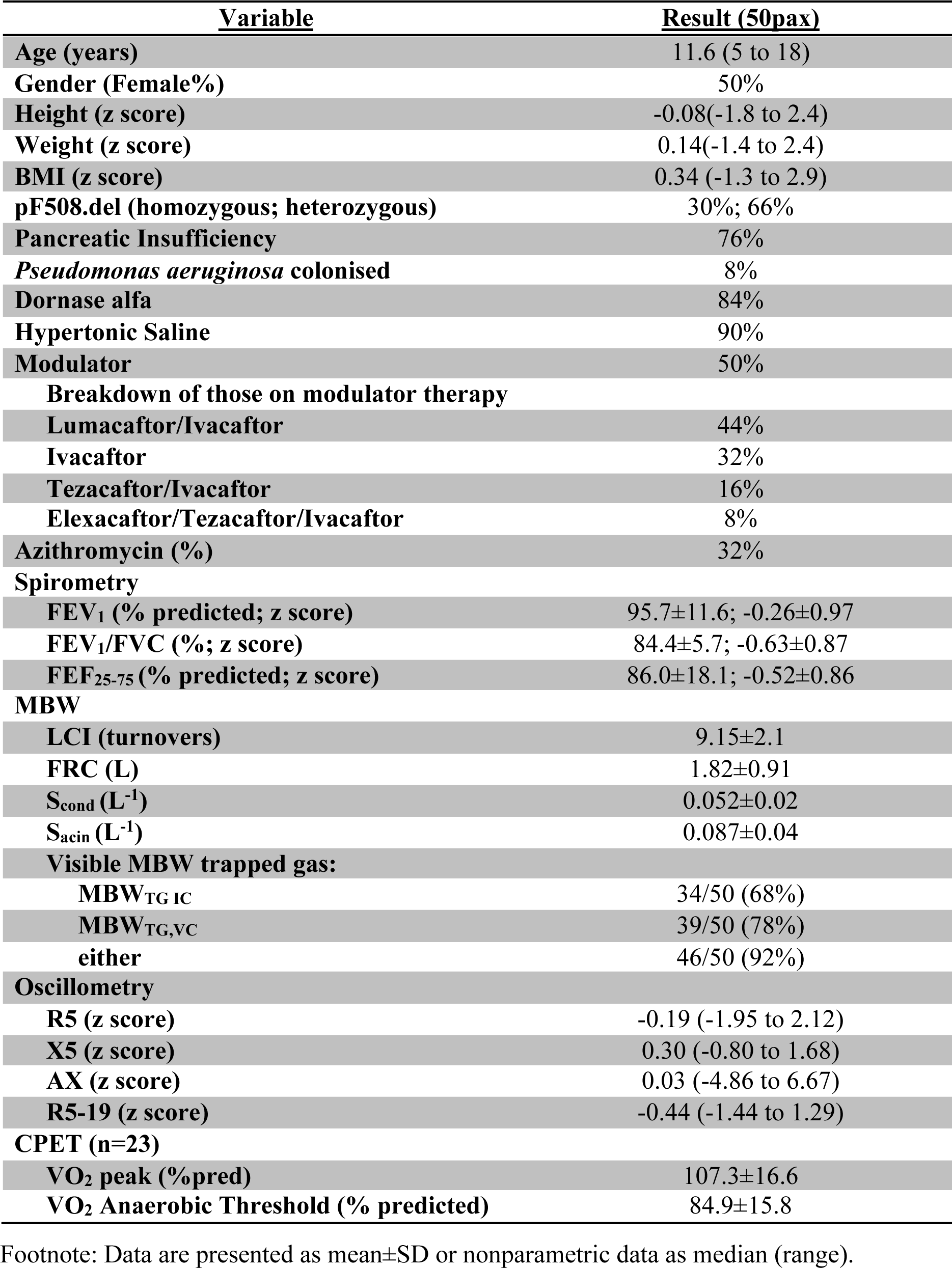
Demographics and Lung Function Characteristics at baseline.

### Rates of structural lung disease

Technical quality of spirometry manoeuvres during CT was good/excellent in 92-100% of scans (Table E1 OLS). Structural disease variables results are summarised in Table E2 OLS. Bronchiectasis, defined as BEI >1, was detected in 24% of patients [BEI 0.57 (0.01 to 10.26)] whilst AT was detectable on all CT scans when defined as any value >0% [AT A_1_ 9.31(0.24 to 38.39)]. Using a 5% threshold, rate of AT detection fell to 58% of patients. This 5% threshold and A_1_ values were used in further analyses (see OLS for justification).

### Lung function

Functional variables at baseline, and rates of abnormality, are summarised in Table 1 and Figure 1, respectively. Of the functional tests, MBW had the highest percentage of patients with defined abnormality: S_cond_ 82%, MBW_TG RV_ 78%, LCI 74%, MBW_TG IC_ 68% and S_acin_ 51%. CPET VO_2_peak detected slightly higher rates of abnormality (9%) than spirometry variables (FEV_1_/FVC 8%, FEF_25-75_ 8%, FEV_1_ 2%). Of the conventional oscillometry variables, AX detected the greatest amount of abnormality (14%), followed by Rrs (2%), whilst R5-19 and Xrs failed to detect any abnormality. Low to moderate correlation was observed between spirometry and MBW (Table E7 OLS): FEV_1_z with LCI (r=-0.52, *p*<0.0001), S_cond_ (r=-0.50, *p*=0.0003) and S_acin_ (r=-0.33, *p*=0.02). Oscillometry did not correlate with MBW, however AX did show weak correlation with FEF_25-75_ (r=-0.37, p=0.015). VO_2_ correlated only with FEV_1_ (r=0.48, *p*=0.02).

**Figure 1:**
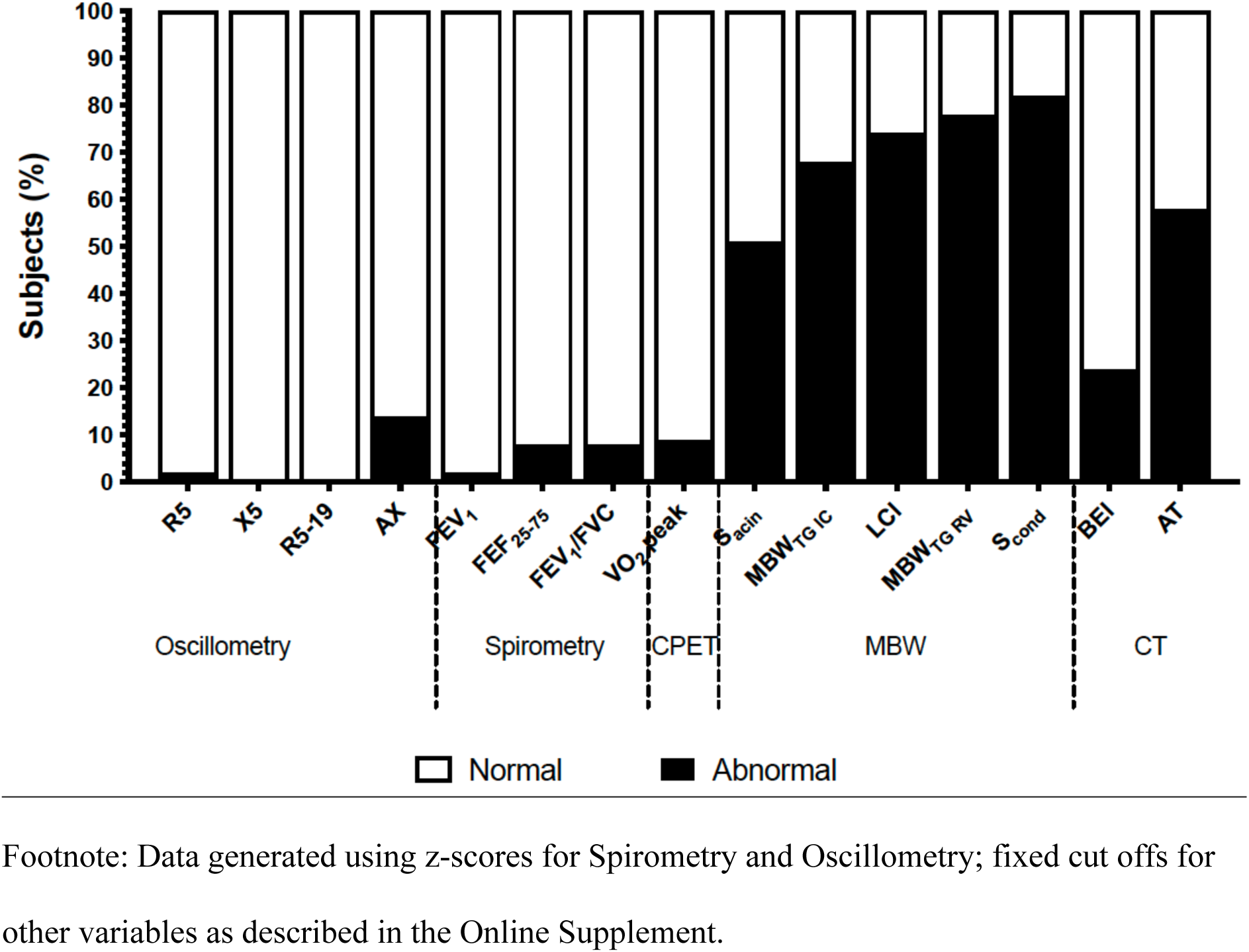
Rates of abnormality across investigated indices of functional and structural lung disease.

### Structure-function relationships

#### Bronchiectasis

Bronchiectasis (as assessed by BEI) correlated with LCI (r=0.46, *p*<0.01; Figures 2 and 3A; Table 2). Bronchiectasis as a predictor of abnormal LCI had perfect sensitivity and negative predictive values (Table 2), with lower specificity and positive predictive values reflecting several subjects with abnormal LCI but no bronchiectasis. S_cond_ but not S_acin_ had similar correlation strength with BEI (Figures 2 and 3C, Table 2). No correlation was observed between spirometry variables and BEI (Figure 2).

**Figure 2:**
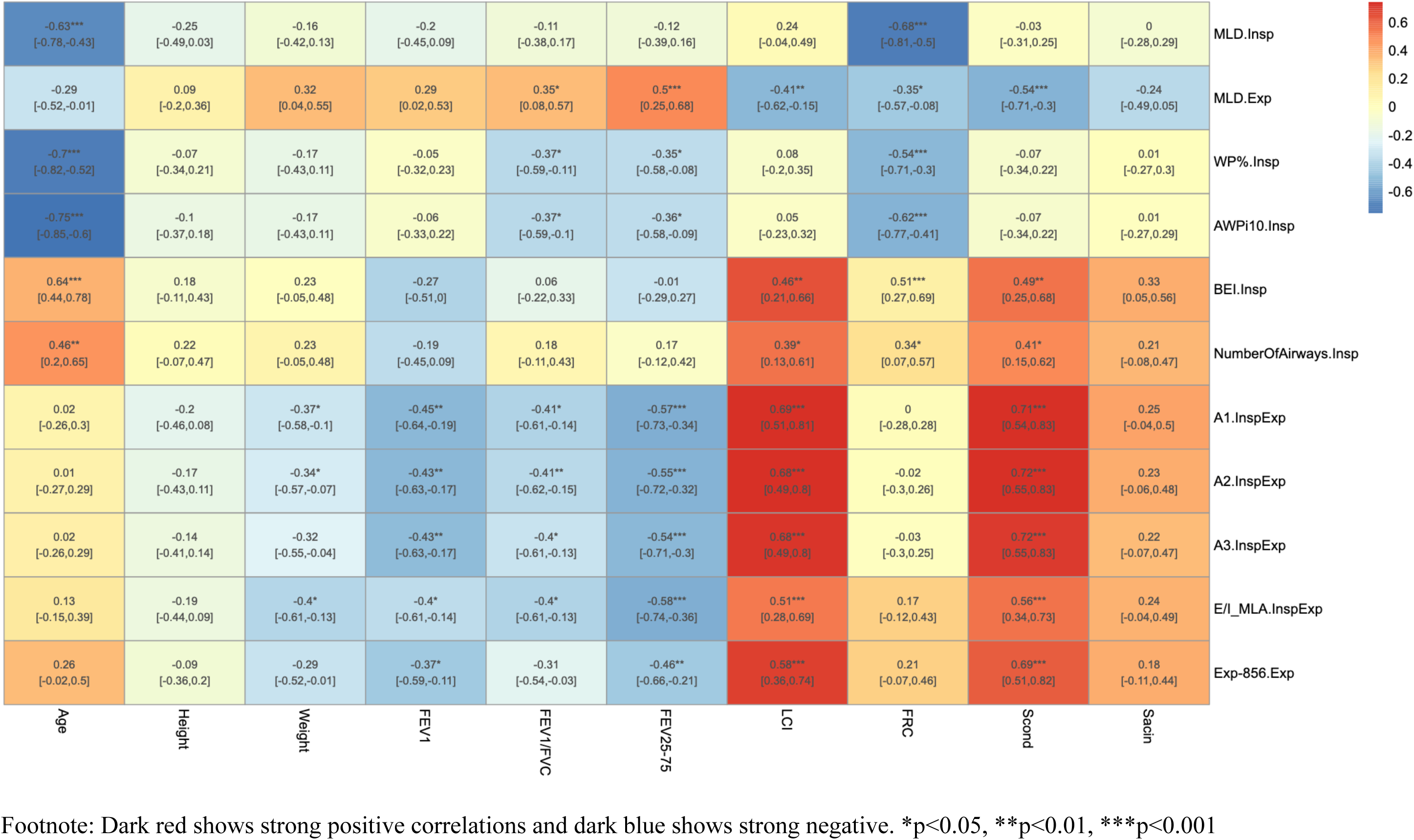
Correlation heat map of structure-function relationships.

**Figure 3.**
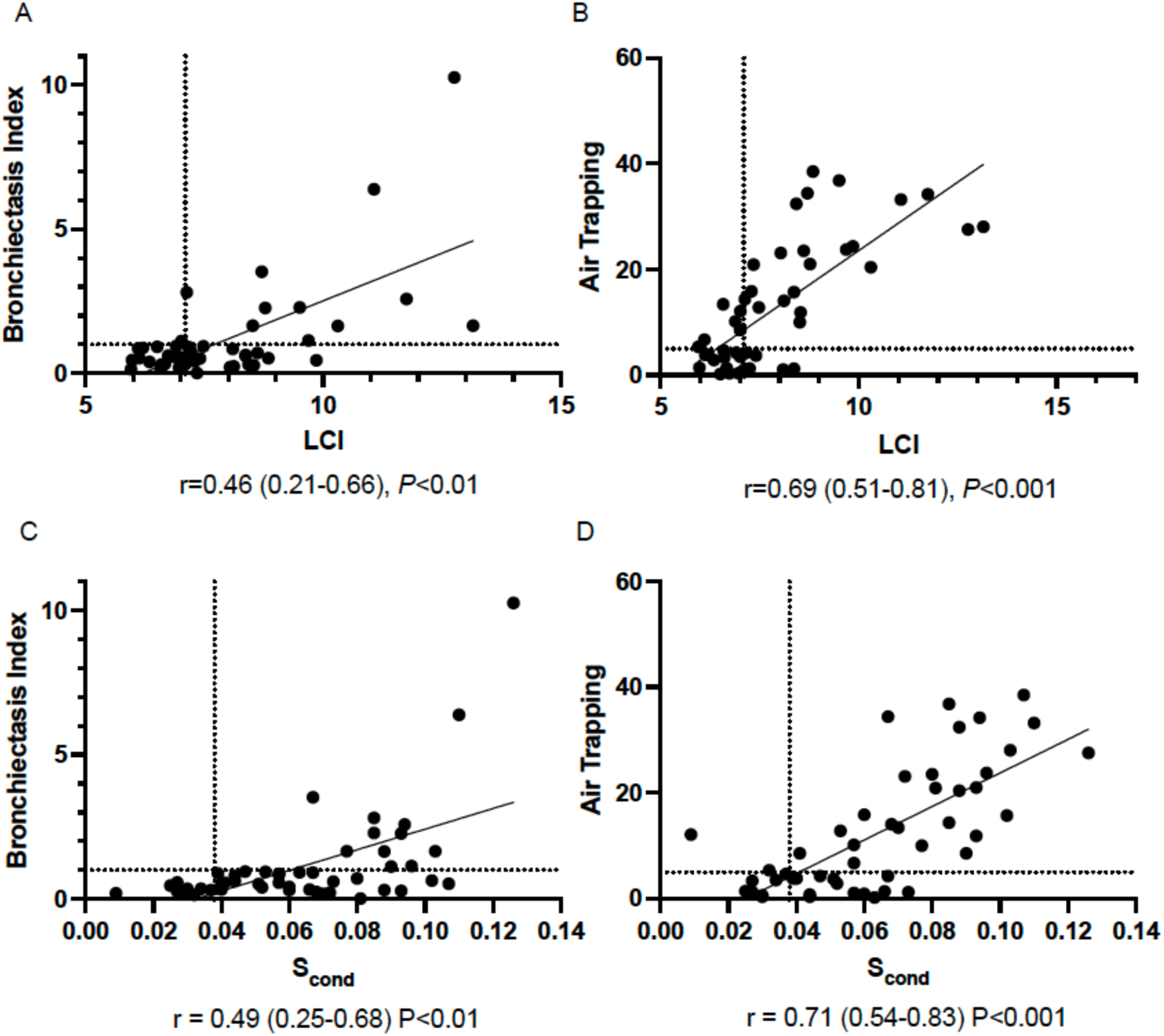
Structure-Function relationships: Bronchiectasis and Air Trapping from CT vs. MBW variables LCI and S_cond_.

**Table 2.** Structure-Function relationships: Bronchiectasis and Air Trapping from CT vs. MBW and spirometry variables.

| **Low dose** |  |
| --- | --- |
| **Bronchiectasis** |  |
| BEI vs. LCI | 0.46 (0.21 to 0.66)** |
| BEI vs. S<sub>cond</sub> | 0.49 (0.25 to 0.68)** |
| BEI vs. S<sub>acin</sub> | 0.33 (0.05 to 0.56) |
| BEI vs. FEV<sub>1</sub> | -0.27 (-0.51 to 0.0) |
| BEI vs. FEV<sub>1</sub>/FVC | 0.06 (-0.22 to 0.33) |
| BEI vs. FEF<sub>25-75</sub> | -0.01 (-0.29 to 0.27) |
| *Bronchiectasis to predict Abnormal LCI (>7.0)* |  |
| **Odds ratio** | N/A |
| **Concordance** | 54% |
| **Kappa coefficient** | 0.24 (0.09 to 0.39) |
| **Sensitivity** | 100 (75 to 100) |
| **Specificity** | 39 (26 to 55) |
| **Positive Predicted Value** | 34 (21 to 51) |
| **Negative Predicted Value** | 100 (80 to 100) |
| **Air Trapping** |  |
| AT vs. LCI | 0.69 (0.51 to 0.81)*** |
| AT vs. S<sub>cond</sub> | 0.71 (0.54 to 0.83)*** |
| AT vs. S<sub>acin</sub> | 0.25 (-0.04 to 0.50) |
| AT vs. FEV<sub>1</sub> | -0.45 (-0.64 to -0.19)** |
| AT vs. FEV<sub>1</sub>/FVC | -0.41 (-0.61 to -0.14)* |
| AT vs. FEF<sub>25-75</sub> | -0.57 (-0.73 to -0.34)*** |
| *Air Trapping to predict Abnormal LCI (>7.0)* |  |
| **Odds ratio** | 6.9 (1.9 to 29.9), P=0.005 |
| **Concordance** | 72% |
| **Kappa coefficient** | 0.40 (0.15 to 0.66) |
| **Sensitivity** | 71 (55 to 84) |
| **Specificity** | 73 (48 to 89) |
| **Positive Predicted Value** | 86 (69 to 95) |
| **Negative Predicted Value** | 52 (32 to 72) |
| **Low dose** |  |
| **Bronchiectasis** |  |
| BEI vs. LCI | 0.46 (0.21-0.66)** |
| BEI vs. S<sub>cond</sub> | 0.49 (0.25-0.68)** |
| BEI vs. S<sub>acin</sub> | 0.33 (0.05-0.66) |
| BEI vs. FEV<sub>1</sub> | -0.27 (-0.55-0.0) |
| BEI vs. FEV<sub>1</sub>/FVC | 0.06 (-0.22-0.33) |
| BEI vs. FEF<sub>25-75</sub> | -0.01 (-0.29-0.27) |
| *Bronchiectasis to predict Abnormal LCI (>7.0)* |  |
| **Odds ratio** | N/A |
| **Concordance** | 54% |
| **Kappa coefficient** | 0.24 (0.09-0.39) |
| **Sensitivity** | 100 (75-100) |
| **Specificity** | 39 (26-55) |
| **Positive Predicted Value** | 34 (21-51) |
| **Negative Predicted Value** | 100 (80-100) |
| **Air Trapping** |  |
| AT vs. LCI | 0.69 (0.51-0.81)*** |
| AT vs. S<sub>cond</sub> | 0.71 (0.54-0.83)*** |
| AT vs. S<sub>acin</sub> | 0.25 (-0.04-0.50) |
| AT vs. FEV<sub>1</sub> | -0.45 (-0.64 to -0.19)** |
| AT vs. FEV<sub>1</sub>/FVC | -0.41 (-0.61 to -0.14)* |
| AT vs. FEF<sub>25-75</sub> | -0.57 (-0.73 to -0.34)*** |
| *Air Trapping to predict Abnormal LCI (>7.0)* |  |
| **Odds ratio** | 6.9 (1.9-29.9), P=0.005 |
| **Concordance** | 72% |
| **Kappa coefficient** | 0.40 (0.15-0.66) |
| **Sensitivity** | 71 (55-84) |
| **Specificity** | 73 (48-89) |
| **Positive Predicted Value** | 86 (69-95) |
| **Negative Predicted Value** | 52 (32-72) |

Footnote: Correlations are presented as Pearson's r, Odds ratio (95% Confidence Intervals)
and Sensitivity, Specificity and Negative Predictive Value in percentage (%). Bronchiectasis Index (BEI) abnormality was defined as >1. Air Trapping abnormality was defined as >5%.
\*p<0.05, \*\*p<0.01, \*\*\*p<0.001

#### Air trapping

In comparison to bronchiectasis, stronger correlations were observed for air trapping with LCI and S_cond,_ (r=0.73-0.74, p<0.001) but weaker with S_acin_ (Figures 2 and 3B/D, Table 2). Air trapping as a predictor of abnormal LCI, had lower sensitivity and negative predictive value, compared to those observed with bronchiectasis, but improved specificity and positive predictive values (Table 2). Trapped gas during IC breaths (MBW_TG IC_) was detected in 68% whilst it was detected in 78% for the additional breath to RV (MBW_TG RV_). Only four subjects (8%) had no evidence of visible MBW TG on either. MBW_TG IC_ and CT air trapping classification was concordant in 74% of subjects (50% abnormal, 24% normal). For discordant cases 18% had abnormal MBW_TG IC_ but normal AT. Equivalent but weaker inverse correlations were observed for CT air trapping with Spirometry (Figure 2).

## DISCUSSION

This is the first study to perform a detailed comparison across an extensive range of physiological tools for early disease detection in an exclusive mild/normal lung disease cohort (defined using spirometry FEV_1_). Performance was assessed in comparison to CT (as a gold standard measure of structural lung disease) collected using spirometry-direction and fully quantitative analysis to optimise structural lung disease detection. We confirmed that the majority of subjects (≈60%) had abnormal air trapping, using an evidence based cut off of 5% to define abnormality, and approximately 25% had bronchiectasis. Across our extensive range of lung function variables, we showed that MBW offered improved sensitivity to other tools such as oscillometry and CPET to detect abnormality and correlate with structural lung disease. Finally, additional utility of MBW may be derived beyond LCI by incorporating S_cond_ and presence of MBW-detected trapped gas. Within our cohort, MBW detected greater rates of abnormality than CT.

Understanding test performance, and calculating minimum sample size for intervention studies, requires data specific to the level of disease severity. We believe the severity of disease studies here is broadly representative of the CF population: Current US CFF registry data outline median FEV_1_ values approaching 100% predicted for 6-19 year oldsm, 90% for adults aged 20-40 years, with 89.1% of pwCF reaching 18 years with normal/mild lung disease on spirometry (i..e, FEV_1_ ≥70%)(37). This dataset advances the historical literature, where studies have typically examined structure-function relationships or test performance in cohorts enriched by pwCF with greater severity disease, as commonly found in the literature. (21, 22, 38, 39). This may over-estimate correlations in true early disease evolution. Using a fully quantitative analysis to structural lung disease definition, and an evidence-based cut off the definition of abnormal air trapping, we describe structure-function relationships for both spirometry and MBW variables but did not find them for oscillometry (based on conventional spectral analysis) or CPET variables (Table 2, Figure 2 and Table E7). The strongest correlations were observed between CT air trapping and LCI or S_cond_ (Figure 2)

CT-assessed air trapping was confirmed as the dominant feature of early structural lung disease, in line with previous studies using CT(5, 19), and present in the majority of subjects. Clinical utility of air trapping detection is supported in the literature by its predictive ability to detect later decline in both structural and functional disease(6, 19). Using a fully quantitative approach to CT analysis we found measurable air trapping in *all* subjects within our cohort, however, the exact threshold to define abnormality lacks consensus agreement. This is important to determine given healthy control data demonstrates some air trapping(40). In addition, there was a pronounced effect of varying the abnormality threshold in our cohort (Table E3). The 5% threshold chosen to define abnormality here was supported by previous CT data from a school-aged healthy control paediatric cohort (n=10) which reported a mean (SD) AT of 4.0±3.1%(41), internally validated by reanalysing the same dataset using a recently developed deep learning convolutional neural network model(42) (4.3±1.6%). In addition, structure-function relationships between AT and LCI appeared optimal when using this 5% threshold (Table E3 and Figure E3).

Characterisation of structure-function relationships in early CF lung disease serves to advance understanding of the optimal approach for detection and surveillance(2, 8). Our data extends current literature by including a greater range of functional tools within the same study and exploring utility of novel indices. We confirm significant associations for both bronchiectasis and air trapping with LCI in school-aged children, with normal FEV_1_(5). Abnormal LCI in our cohort increased probability of air trapping on CT. Based on the 5% threshold we used to define abnormal air trapping on CT, the highest rates of abnormality were in MBW, not CT, indices. S_cond_ showed higher rates of abnormality than LCI. This is consistent with results from a mixed paediatric/adult cohort. Within this Horsely et al study, which included pwCF with FEV_1_ values as low as 29% predicted, the additional utility of S_cond_ to LCI within CF was questioned due to an apparent ceiling value for S_cond_ of 0.150, which resulted in no correlation between LCI and S_cond_ in their cohort(39). However, the primary utility of MBW lies within early lung disease(8), and by focusing on that setting within our cohort we clearly show that S_cond_ values are far below that apparent ceiling (all less than 0.100 in our cohort) and strong correlations were observed between S_cond_ and LCI (r=0.85, p<0.001) and between S_cond_ and indices of structural disease (e.g. AT and BEI). Interestingly, S_acin_, along with LCI, was the strongest predictor of bronchiectasis in our cohort, suggesting S_acin_ becomes abnormal later with more advanced disease as suggested by others(39). AREST CF study data, using semi-quantitative PRAGMA analysis, described the negative predictive value (NPV) of LCI for bronchiectasis in school-aged children to be 55%(43). In our cohort, using fully quantitative CT analysis, NPV was higher at 92%, with only one subject having a normal LCI but evidence of bronchiectasis. The complementary nature of CT and functional indices was evidenced by the fact that of those with a normal BEI value (i.e. no bronchiectasis) 24/36 (66%) had an abnormal LCI. The fact that 13/24 (54%) of subjects with abnormal LCI had abnormal AT highlights the role that AT may play in driving early LCI abnormality. Examining imaging and MBW-based assessment of trapped gas, discordancy occurred in 18% where subjects had abnormal MBW_TG IC_ but normal AT on CT.

We further explored MBW’s ability to detect air trapping in this setting. Using a series of IC breaths to detect trapped gas in CF subjects was first described by Gustafsson et al almost 30 years ago(43), and in our early disease cohort visible trapped gas during the 5 IC breaths (MBW_TG IC_) was detectable in two thirds of subjects. We added an additional breath down to residual volume to assess additional trapped gas release during expiration between FRC and RV (MBW_TG RV_) which was detectable in 78% of subjects. No MBW-detectable gas trapping occurred in only 8% of subjects. Strongest predictors of MBW_TG IC_ were LCI and BEI. MBW TG measurements appear complementary, providing differing information about TG release when breathing to different lung volumes i.e. to FRC for MBW_TG IC_ and to RV for MBW_TG RV_. Given that CT estimates of air trapping reflect residual trapped air present after reaching RV, relationships between these measures of air trapping at these different lung volumes should be explored further. Separate work by our group has shown that this MBW trapped gas assessment is feasible down to the young pre-school age range in pwCF(44). Future work will need to outline the clinical impact of MBW-detected trapped gas, but it is interesting to see that dynamic hyperinflation (or increase in gas trapping) during CPET has been shown to be associated with a greater rate of FEV_1_ decline over the subsequent four years(45).

With FEV_1_ values ≥70% predicted, the low rate of abnormality for FEV_1_ (2%) was not surprising but rates of detected abnormality remained poor for other potentially more sensitive spirometric measures: 8% for FEF_25-75_ and 8% for FEV_1_/FVC. CPET and oscillometry are other functional tests proposed as more sensitive measures than spirometry, but in this setting did not perform significantly better. Peak VO_2_ was rarely abnormal on CPET testing. In cohorts containing a range of disease severity, authors have reported peak VO_2_ as having the strongest correlation with measures of structural lung disease (46) and ventilation inhomogeneity(47). However, specific to this early disease setting we did not find correlations between VO_2peak_ or VO_2AT_ and either structural or functional disease variables. Previous studies of impulse oscillometry (IOS) in CF have suggested correlation with LCI(48), but have failed to provide consistent evidence of benefit over spirometry(49, 50). Within our cohort conventional oscillometry variables (Rrs5, Xrs5, AX and R5-19 expressed as a mean value over recordings) appeared to have limited utility: AX-detected abnormality performed slightly better than spirometry variables, but there was no correlation with structural lung disease and only AX showed any correlation (albeit weak) with any other functional variable, which was FEF_25-75_. No correlation with MBW was observed. Although there was a recent publication outlining correlations between LCI and IOS variables(48), our lack of correlation is consistent with previously published AREST CF data in the younger preschool age range(51).

Study strengths include performance of low dose CT using the gold standard spirometer-directed technique performed to a high standard by the recruited subjects; this approach has been shown to optimise air trapping detection on expiratory scans(12), and is recommended to ensure volume standardisation on both inspiratory and expiratory scans(13). School-aged children were intentionally recruited as a cohort where spirometry directed CT could be performed to a high standard to optimise the gold-standard measure of early lung disease used in this study (see OLS for discussion of experience in younger children). As evidence of this strong data quality, with no difference in the spirometry-manoeuvre acceptability (Table E6 OLS) or structure-function relationships (Figure E5 OLS) was found between younger and older age groups (<10years & >10years). The extensive range of functional tests and novel analyses performed allowed more detailed comparisons of sensitivity and specificity than in existing literature. Limitations include lack of strong normative data for some imaging and functional indices (e.g. air trapping on CT and trapped gas on MBW). Whilst the AT threshold chosen was based on both normative data and structure-function relationships present, future work must validate this threshold. Rates of discordance between methods for AT detection will be important to re-assess as more robust thresholds are developed. Finally, this comparison was based on cross-sectional data and longitudinal measurements will be collected to compare ability to detect change over time.

In conclusion, using fully quantitative CT analysis and an extensive array of lung function variables (both conventional and novel), we have demonstrated not only significant structural and functional deficits in early CF lung disease, but significant structure-function relationships which aid future efforts to optimise early disease detection. High rates of abnormality were detected by MBW and CT indices, and for MBW, additional utility, beyond that offered by LCI, was suggested for S_cond_ and MBW-based assessment of gas trapping. Whilst future work will target further evidence of the correct thresholds for defining abnormality, the evidence provided by our study reinforces the complementary nature of these tests and the limited utility of conventional approaches to oscillometry and CPET.

## Supporting information

Online supplement Bayfield et al

## Data Availability

All data produced in the present study are available upon reasonable request to the authors

## ACKNOWLEDGMENTS

Authors would like to thank the Children and Families who took part in this study, staff of the Respiratory Function Unit and Radiology Department that assisted during the study, SCHN Ethics and Governance and Kids Research at CHW. A 2018 Australian CF Research Trust Innovation Grant funded this study.

## CONFLICT OF INTEREST STATEMENT

The authors have no conflict of interests to declare

## Notes

### Competing Interest Statement

The authors have declared no competing interest.

### Author Declarations

Ethics committee approval from the Sydney Children's Hospitals Network Human Research Ethics Committee (SCHN HREC, ethics approval 18/SCHN/469) and written informed consent/assent was obtained from all parents/guardians and children

