## Supplementary material for "Comparative sensitivity of Early Cystic Fibrosis Lung Disease detection tools in school aged children": Online supplement Bayfield et al

*Online Data Supplement*

### SUPPLEMENTAL METHODS

The aim was to perform low dose CT and all other tests on the same day, which was achieved in most cases (n=43/57 subjects, 75%). In all other cases tests were performed within two weeks. In those where testing occurred on different days, this was due to CT slot availability or lung function testing being performed outside of CF clinic slots.

#### Multiple breath washout (MBW)

*Quality control*

Technical acceptability of MBW trials was assessed using published quality control criteria[1], based on ERS/ATS inert gas washout consensus statement recommendations[2]. Concentration normalized phase III slope ($Sn_{III}$) analysis involved visual inspection and estimation of phase III slopes ($S_{III}$) for each breath, with $S_{cond}$ and $S_{acin}$ calculated per trial and averaged across triplicate trials as per the Spiroware software (v3.1.6) used for data collection. Only trials with acceptable $S_{III}$ for the first breath of each trial and at least two thirds of breaths between 1.5-6.0 lung turnovers were accepted. Due to the description recently of a cross-talk error between the oxygen ($O_2$) and Carbon dioxide ($CO_2$) analyzers[3 4], and the availability of updated commercial software to correct for this (v3.3.1), all MBW data was updated by rerunning the original A-files in this latest version. Reference values to define abnormality for LCI, FRC and $Sn_{III}$ variables used were equipment and software version specific[5] and had also been corrected for this error (personal communication with EcoMedics AG). The presence of trapped gas was defined as any increase in end tidal $N_2$ concentration, compared to the tidal breath immediately prior to the VTG breathing manoeuvres. In a separate cohort of n=10 school-aged healthy control subjects we confirmed that end-tidal $N_2$ values continue to decrease through this VTG breathing protocol in health.

### Spirometer-directed computed tomography scanning protocol

*Spirometry-directed manoeuvre acceptability criteria*

Inspiratory and expiratory standard low dose CT scans are taken routinely within the CHW CF clinic at three points during childhood (approximately at age 5-7, age 10-12 and age 15-17 years). All of the children who took part in this study were due their clinical surveillance CT scan. The expiratory scans were taken during a breath-hold at residual volume (RV), after a full exhalation, and the inspiratory scans were taken during a breath-hold at Total Lung Capacity (TLC) after inspiration from RV (Supplemental Figure E1). Supine spirometry vital capacity (VC) values were compared to supine practice values ("target value") and graded using published criteria[6]: for inspiratory scans, <85% of target value= suboptimal, ≥85-95 = good, and ≥95% = excellent; for expiratory scans, <80% of target = suboptimal, ≥80-90 = good, and ≥90% = excellent. Participants <10 years had at least one additional supine spirometry practice visit before the CT scan, typically completed at the time of a scheduled outpatient visit for clinical care and only when routine seated spirometry was already deemed acceptable and repeatable. Only children with acceptable and repeatable seated & supine spirometry were deemed eligible to be approached for recruitment into the study.

**Figure E1:** Volume time graph depicting the breathing phases for spirometry-directed CT. Expiratory scans were taken during a breath-hold at residual volume (RV), after a full exhalation from total lung capacity (TLC), and the inspiratory scans were taken during a breath-hold at TLC after inspiration from RV. Separate manouevres were performed for each scan.

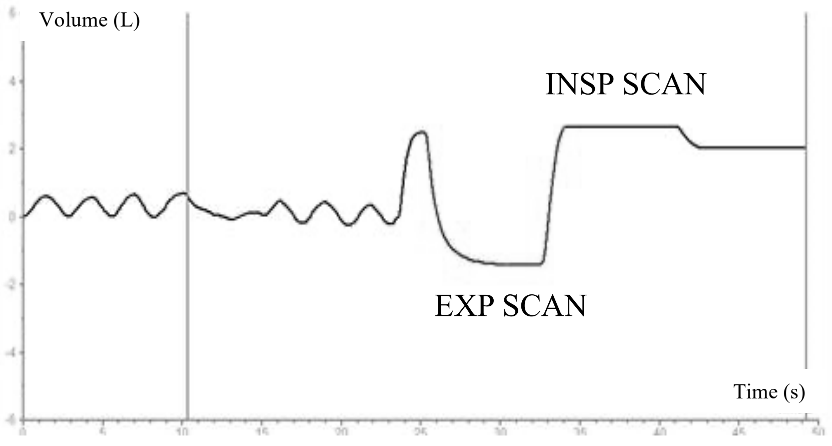


*CT scanning Dose configuration*

The LD scanning technique was specified as follows: reference 100 kV with tin filter, 192x0.6 mm detector configuration (96x0.6 mm physical detector collimation with a z-flying focal spot), 35 quality reference mAs for inspiratory and 150mAs for expiratory, 0.25 second rotation time for inspiratory and 0.5 second rotation for expiratory, and 0.6 helical pitch for inspiratory and 1.2 for expiratory. The reconstruction settings used were Kernel Br49d\3, slice thickness 0.6mm and slice gap 0.3mm.

*Radiation exposure*

Effective CT radiation dose was calculated using a chest conversion factor[7] following updated guidelines[8], the main manuscript shows mSv results from all scans i.e. TOPO, inspiratory and expiratory. CT dose index (CTDI) for inspiratory scans was median 0.7 (range 0.24-

2.08)mGy, whilst for expiratory scans CTDI was 0.37 (0.18-0.82)mGy. Effective dose for paired inspiratory and expiratory CT was 0.66mSv (0.33-2.0), whilst SSDE was 1.12 (1.14-3.31) and 0.58 (0.36-1.62), respectively. Effective radiation dose for LD inspiratory and expiratory scans was 0.46 (0.20-1.46) and 0.20 (0.12-0.56) mSv, respectively.

*Quantitative CT post-processing*

The fully automated YACTA software (version 2.9.1.12) initially performed segmentation and lobe-based labelling of the airways and the lungs on the paired inspiratory and expiratory CT images[9-12]. Then quantitative CT (QCT) variables were determined.

QCT variables for the airways were calculated on the inspiratory scans. The voxel-based result of the airway tree segmentation was skeletonized and transformed to an acyclic graph. An anatomical knowledge-based algorithm labelled central airways and lobar bronchi. The direction vector of an airway, which can be determined by the graph representation of the airway tree, was used to calculate orthogonal planes for every airway. On these planes the airways were assessed lobe- and generation-based for lumen area (LA), wall thickness (WT), total diameter (TD) and wall percentage (WP) using the parameter-free integral-based method[13-17]. The square root of the wall area for a 'theoretical airway' with an internal perimeter of 10 mm (AWT-Pi10) was determined as described by Grydeland *et al.*[18]. A bronchiectasis index (BEI) was calculated as described by Weinheimer *et al.*[16].

QCT variables for the lung parenchyma were determined by using the inspiratory and expiratory scans. In this study the focus was on the air trapping (AT) quantification. AT was quantified by 3 different variables:

1. E/I MLA which is the expiratory to inspiratory ratio of mean lung attenuation with a range from 0 to 1.0, greater values mean more air trapping [19]. This has the disadvantage of not adjusting for the normal changes in lung density that occur with age[20];
2. Determination of A1, A2 and A3 which uses three thresholds for the definition of air-trapping, expressing the size of the defect areas as a fraction of the total segmented expiratory lung. A1 represents defects on the basis of liberal criteria, which includes mild, moderate, and severe air trapped regions, while A3 represents defects on the basis of stringent criteria as published previously (severe air-trapping)[21]. This more adaptive approach takes into account lung inflation. We used the A1 parameter introduced in Goris et al for air trapping quantification[21]. A1 indicates the percentage of mild, moderate, and severe air trapping throughout the whole lungs. A1 is based on a patient specific threshold T1 determined on the basis of the expiratory and inspiratory histograms of the lungs. This threshold T1 is applied to the expiratory CT images. Each air trapping segmentation was visually inspected. We have previously used and validated this automated segmentation approach in the following publications[9 14 22].
3. "Exp-856.Exp" as the most common air trapping parameter used in adult studies[19].

**Classification of Abnormality**

For spirometry, z score values outside ±1.96 were considered abnormal for $FEV_1$, $FEV_1/FVC$ and $FEF_{25-75}$[23-25]. For MBW the published upper limits of normal (ULN) specific to the ExhalyserD were used: Lung clearance index (LCI) >7.1[26], $S_{cond}$ abnormal >0.038, $S_{acin}$ abnormal >0.073 (direct communication with EcoMedics AG). LCI ULN for children aged 3-6 years was found to be >7 from published preschool cohorts[27] reanalysed in v3.3.1 (personal communication with Felix Ratjen). For oscillometry, z score values outside ±1.96 were considered abnormal for R5, X5, R5-19 and AX based on equipment specific reference equations[28].

For CT imaging analysis, Bronchiectasis Index (BEI) abnormality was defined as >1. For AT, of the three previously developed thresholds (A1, A2 and A3), A1 was selected due to better performance in previous CF CT studies[29 30]. As explained in the main manuscript a 5% threshold for abnormality was chosen based on two main factors. Firstly previous authors within the collaborative group (TE, CG) had performed modelling with a limited chest CT dataset that utilized spirometer-controlled chest CT imaging in n=10 normal paediatric subjects recruited for a CF vs. healthy controls pilot study[31]. In this study the mean (range) AT value was 4.0±3.5%, based on six expiratory slices and a quantitative threshold technique (25 delta Hounsfield Units assessment)[31]. Secondly, the better structure-function relationships observed at this threshold (see later sections in OLS).

### SUPPLEMENTAL RESULTS

#### Quality control of Spirometry manoeuvres during CT scanning

Inspiratory and expiratory scans were classified as described earlier (Table E1).

**Table E1:** Spirometer directed grading classification for both Inspiratory and Expiratory CT scans.

| | Insp LD | Exp LD |
|---|---|---|
| **Excellent** | 70 | 96 |
| **Good** | 22 | 4 |
| **Suboptimal** | 8 | 0 |

Footnote: Classification results are based on the percentage difference between the supine spirometry practice $FEV_1$ values and actual spirometry values during the CT scan image acquisition[6]. Values shown are the percentage of scans fulfilling each grading classification.

**CT Quantification performance for LD CT scan analysis**

YACTA is well-evaluated, fully automatic CT analysis software. Average time to complete analysis was ~20 minutes. In this cohort there were approximately 815 lung slices per scan (inspiration: 880, expiration: 750). Segmentation of airways was achieved in all scans, detecting 132.5 (33 to 424) airways. Results obtained are summarised in Table E2.

**Table E2:** CT analysis from the automated quantitative YACTA platform.

| CT indices | Low Dose |
|---|---|
| **Air Trapping** | 9.31(0.24 to 38.39) |
| **Bronchiectasis Index (BEI)** | 0.57 (0.01-10.26) |
| **Wall Percentage** | 51.63±5.87 |
| **Wall Thickness** | 1.26±0.14 |
| **Total Diameter** | 8.04±0.81 |
| **Lumen Area** | 31.58 (18.78 to 55.61) |
| **Mean Lung Density** | -502.4±64.24 |
| **Average Number of Airways** | 132.5 (33 to 424) |
| **CTDIvol Insp** | 0.76 (0.21 to 2.08) |
| **CTDIvol Exp** | 0.36 (0.08 to 0.82) |
| **SSDE Insp** | 1.12 (0.38 to 3.31) |
| **SSDE Exp** | 0.58 (0.15 to 1.62) |
| **DLP Insp** | 21.39 (3.40 to 54.32) |
| **DLP Exp** | 8.17 (1.82 to 18.2) |
| **Effective diameter Insp** | 23.35 (17.31 to 30.97) |
| **Effective diameter Exp** | 20.7 (16.2 to 27.7) |
| **Tracheal Air HU Insp** | -972.2 (-982.0 to -943.3) |
| **Tracheal Air HU Exp** | -953.0 (-987.5 to -876.5) |
| **Tracheal Air SD Insp** | 27.1 (19.8 to 41.6) |
| **Tracheal Air SD Exp** | 43.9 (24.6 to 98.5) |
| **Insp Lung Volume (ml)** | 3105 (1593 to 7460) |
| **Exp Lung Volume (ml)** | 999.4 (431.8 to 2667) |
| **Average Measurements available per Generation** | |
| **G1** | 165.4±39.2 |
| **G2** | 119.5 (19 .0 to 201.0) |

| | |
|---|---|
| **G3** | 37 (15.0 to 67.0) |
| **G4** | 93.0 (50.0 to 175.0) |
| **G5** | 152.5±48.2 |
| **G6** | 199.3±95.6 |
| **G7** | 196.5 (16.0 to 584.0) |
| **G8** | 160.0 (9.0 to 699.0) |
| **G9** | 86.5 (3.0 to 656.0) |
| **G10** | 37.0 (1.0 to 460.0) |

Footnote: Data shown as median (range) or mean ± standard deviation

**Structure Function relationship**

*Air Trapping*

Quantitative air trapping thresholds have been used previously in the literature[21 29 30]. Correlation of the three thresholds [A1, A2 and A3][21 29 30] with LCI are summarised in Figure E2. Air Trapping threshold AT (A1) represents all air trapped regions, while AT A2 represents a mild-moderate trapped gas (TG) threshold and AT A3 represents a severe TG threshold. Those with a higher LCI have more AT in all panels and all AT results have moderate/strong correlations.

All three thresholds had strong correlation with LCI but as A1 describes AT from all regions of the lung and this threshold is similar to indexes presented by Bonnel *et al.* 2004[31], A1 was utilised for our main AT results and below. Unless specified otherwise AT results are generated using the A1 parameter.

AT was detectable on all LD CT scans when defined as any value > 0%. Therefore, different AT abnormality thresholds were compared (1%, 5% and 10%). The proportions with air trapping decreased as the threshold value increased (Table E3). Factors determining the final decision to use a 5% AT threshold to define abnormality are now described. ROC analysis was performed to examine the ability of LCI to detect air trapping based on AT (A1) at these 1%, 5% and 10% thresholds (Table E3, Figure E3. Strong AUC values (≥0.80) and sensitivity were observed using a AT threshold of ≥5% to define abnormality, with equivalent specificity was observed with 5% and 10% thresholds.

**Figure E2:** Correlation plots of quantitative Air Trapping thresholds $A_1$, $A_2$ and $A_3$ in CT scans vs. LCI.

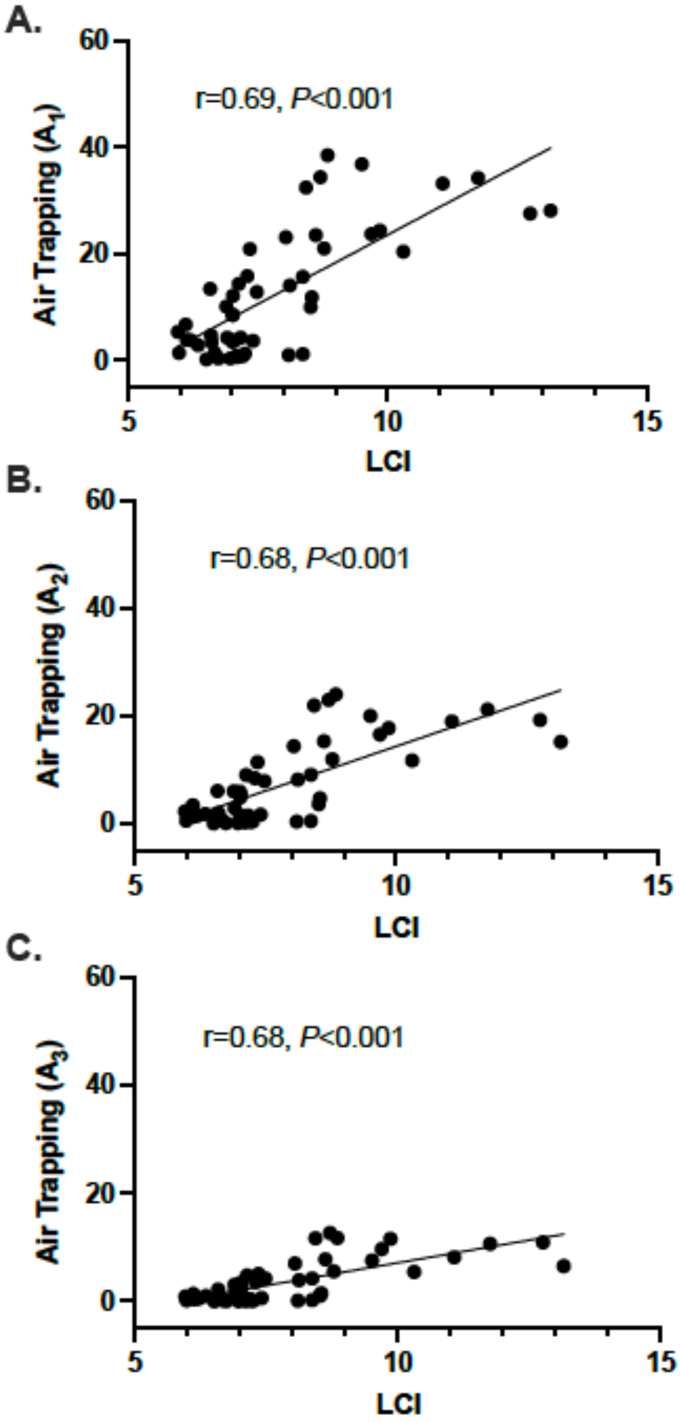


**Table E3:** Air Trapping threshold analysis. Sensitivity and Specificity of abnormal LCI (>7.0 turnovers).

| | 1% | 5% | 10% |
|---|---|---|---|

| | | | |
|---|---|---|---|
| **% of patients with AT** | 88% | 58% | 50% |
| **% of patients with no AT** | 12% | 42% | 50% |
| **AUC** | 0.69;*P*=0.12 | 0.81;*P*=0.06 | 0.91; *P*=0.04 |
| **Sensitivity** | 73 (58-84) | 86 (69-94) | 92 (75-99) |
| **Specificity** | 50 (19-81) | 52 (32-72) | 52 (34-70) |

**Figure E3:** Receiver Operator Curves (ROC) with three different trapped gas (TG) thresholds (1%, 5% and 10%) for AT in low dose CT scans to determine the sensitivity and specificity of lung clearance index (LCI) to detect gas trapping.

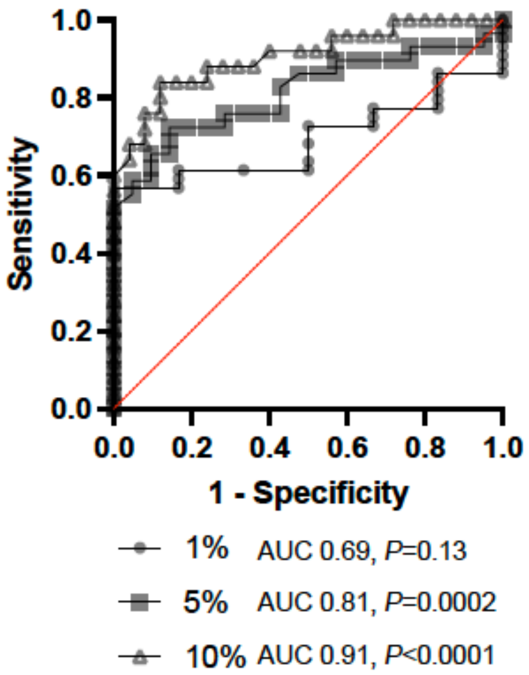


Data from a school-aged healthy control paediatric cohort (n=10) had a mean (SD) AT of 4.0±3.5%, based on 6 expiratory slices and a quantitative threshold technique (25 delta Hounsfield Units assessment)[31]. Internal validation of this threshold using the same dataset and a recently validated convolutional neural network model[32] obtained similar values, 4.3±1.6%, suggesting the AT threshold chosen of 5% was suitable.

The justification to include E/I MLA results was strengthened by the fact that E/I MLA values did not display the same offset observed for A1. The correlations observed for both E/I MLA and Exp-856.Exp were weaker than A1-3.

Concordance in classification of abnormality for LCI in those with and without air trapping on low dose CT imaging is summarized in Table E4.

**Table E4.** Concordance in classification of abnormality for LCI in those with and without air trapping (AT>5%) on low dose CT imaging.

| | | **Low dose CT air trapping** | | |
|---|---|---|---|---|
| | | **Positive** | **Negative** | **total** |
| **Abnormal LCI** | **Positive** | 25 | 10 | 35 |
| | **Negative** | 4 | 11 | 15 |
| | **total** | 29 | 21 | 50 |

*Bronchiectasis Index*

Concordance in classification of abnormality for LCI in those with and without bronchiectasis on low dose CT imaging is summarized in Table E5.

**Table E5.** Concordance in classification of abnormality for LCI in those with and without bronchiectasis (BEI>1) on low dose CT imaging.

| | | **Low dose CT bronchiectasis** |
|---|---|---|

| | | Positive | Negative | total |
|---|---|---|---|---|
| Abnormal LCI | Positive | 12 | 23 | 35 |
| | Negative | 0 | 15 | 15 |
| | total | 12 | 38 | 50 |

BEI showed moderate correlations with LCI and $S_{cond}$ and low correlation with $S_{acin}$. A strong ability (AUC>0.80) was observed for LCI to predict bronchiectasis (Figure E4).

**Figure E4:** Receiver Operator Curves (ROC) for lung clearance index (LCI) to determine the sensitivity and specificity to detect Bronchiectasis (defined as BEI>1) in low dose CT scans.

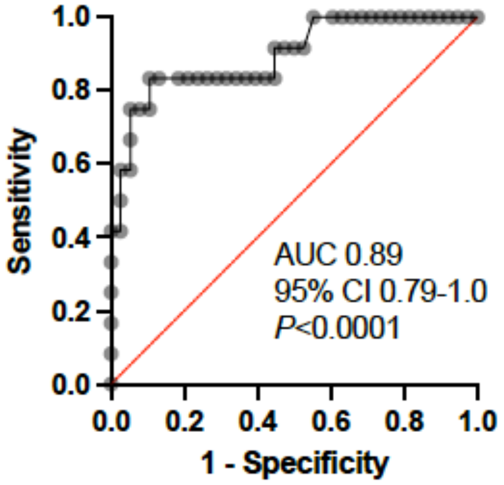


Wall percentage during inspiration, or "WPInsp", was not a sensitive index and correlated poorly with functional indices. This was not unexpected. When higher values are seen it reflects bronchial wall thickening and is a measure for the initial changes in airway remodelling. This parameter is not felt to work as well once bronchiectasis is present as the wall lumen becomes larger.

**Age effects on structure-function relationships**

To look for age effects, the results obtained in children aged ≥10 years (n=32) were compared to those obtained in children aged <10 years (n=18). During CT data acquisition, there were no significant differences between age groups in spirometry volumes as percentage of supine practice values (Table E6). This provides confidence that the spirometer directed technique was highly feasible and performed to the same high standard in our younger patients.

**Table E6:** Percentage difference between practice and scan FVC values by age category (<10 years, ≥10 years) for both Inspiratory and Expiratory CT scans.

| | <10years (n=18) | ≥10years (n=32) | *P* value |
|---|---|---|---|
| **Inspiratory** | -1.37±10.48 | 0.12±8.55 | 0.59 |
| **Expiratory** | 0.35(-13.1 to 10.5) | 0.97(-7.8 to 12.9) | 0.62 |

Footnote: Results are expressed as the percentage difference between supine spirometry practice FVC values (target value) and actual spirometry values during the CT scan image acquisition.

When comparing the lung function characteristics of the younger age group to the older age group (<10years vs. ≥10years), there were no significant differences between $FEV_1$ and $FEF_{25-75}$ (-0.07±0.87 vs. -0.36±1.0, *p*=0.3 & -0.58±0.84 vs. -0.48±0.91, *p*=0.7). LCI was numerically lower in the younger age group but did not reach statistical significance: 7.18 (5.96 to 11.75) vs. 7.31(5.98 to 13.15), *p*=0.51.

Structure-function relationships were maintained across the two age groups. Correlations and correlation slopes were similar comparing younger and older age groups, for both AT and BEI plotted against either LCI (Figure E5) or $FEV_1$ (data not shown).

**Figure E5:** Low dose (LD) CT scan assessed Air Trapping and Bronchiectasis Index scores vs. LCI for those under 10 years (n=18) and those aged 10 years and above (n=32).

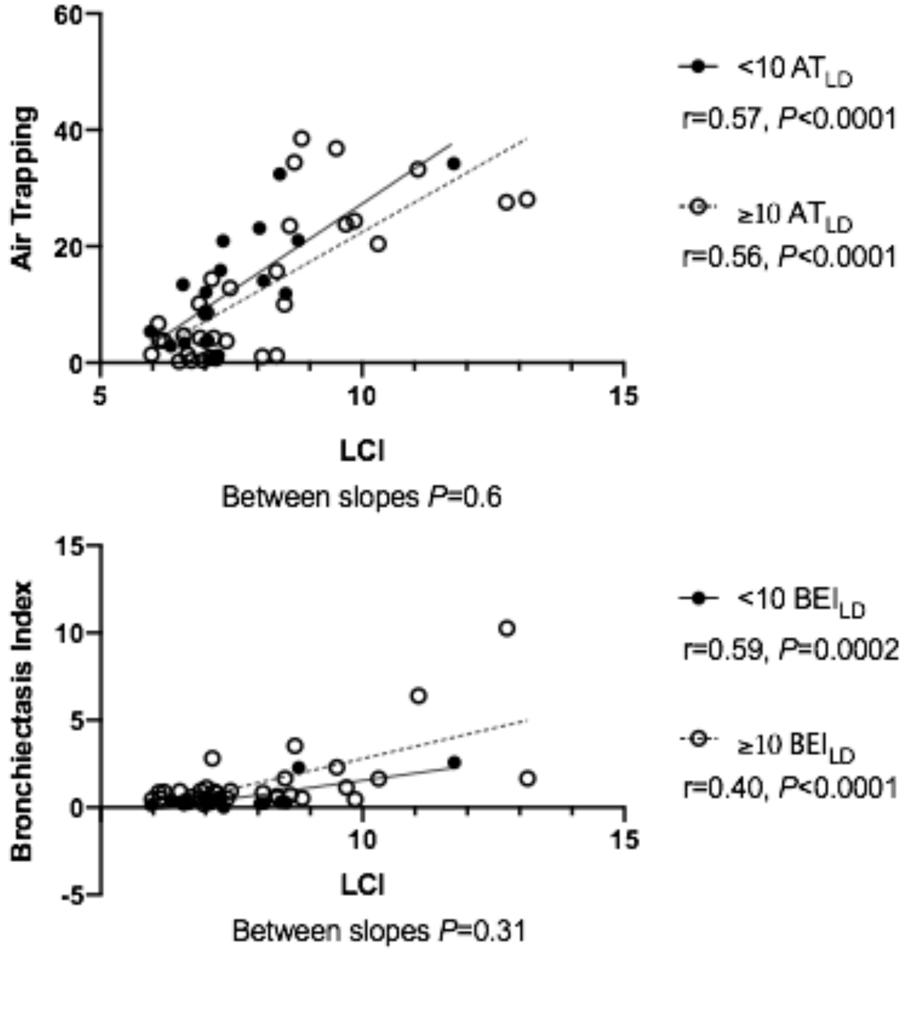

The proportion of AT abnormality did not differ between the age groups when defined as AT>5%. For AT, 53% of participants ≥10years had abnormal AT (17/32) compared to 67% (12/18) of those <10years (*p*=0.39). BEI was lower in the younger cohort: 0.32(0.01 to 2.58) vs. 0.86(0.19 to 10.26), *p*<0.0001. When bronchiectasis was defined as BEI >1; 37.5% had abnormal BEI in those ≥10years and 11% in <10years (*p*<0.0001).

Our decision to impose the lower age limit we did was supported by our experience in preparing the younger school age children for the study. We only approached those with documented evidence of reproducible spirometry across ≥3 clinic visits, yet in many of those aged 5-6 years, fulfilling this criterion, we found a significant variability in supine spirometry values and marked differences between sitting and supine values. We believe this would have affected the accuracy of bronchiectasis detection and quantitative air trapping estimates. Defining the optimal training strategy for these younger children and the thresholds for volume reproducibility on key CT outcomes will be an important focus of future work.

**Clinical measures**

Clinical surrogates of increased CF disease severity i.e. pancreatic insufficiency and increased prescription of medications were related to both functional and structural outcomes, supporting clinical relevance of these indices (Table E7). Abnormal LCI was associated with pancreatic insufficiency, azithromycin use and greater IV antibiotic use (total days) in the preceding year. Dornase alfa prescription was associated with abnormal LCI values and lower $FEV_1$. Those prescribed Azithromycin had increased BEI and total IV antibiotic days in the preceding year positively correlated with BEI.

**Table E7**: Correlation and Area under the curve (AUC) results between functional measures, CT measures of structural disease and clinical measures.

| | $FEV_1$ | $FEV_1/FVC$ | $FEF_{25-75}$ | LCI | $S_{cond}$ | $S_{acin}$ | $BEI_{LD}$ | $AT_{LD}$ |
|---|---|---|---|---|---|---|---|---|
| **Correlations** | | | | | | | | |
| **$FEV_1z$** | - | 0.12 | 0.56*** | -0.52*** | -0.50** | -0.33* | -0.40** | -0.52*** |
| **$FEV_1/FVC$** | - | - | 0.78*** | -0.36* | -0.37** | -0.42** | -0.19 | -0.40** |
| **$FEV_{25-75}$** | - | - | - | -0.52*** | -0.49*** | -0.39** | -0.33* | -0.55*** |
| **LCI** | - | - | - | - | 0.85*** | 0.62*** | 0.64*** | 0.79*** |
| **$S_{cond}$** | - | - | - | - | - | 0.51*** | 0.51*** | 0.74*** |
| **$S_{acin}$** | - | - | - | - | - | - | 0.43** | 0.43** |
| **$MBW_{VTG}$** | -0.20 | -0.20 | -0.22 | 0.4** | 0.36* | -0.01 | 0.30* | 0.44** |
| **$VO_{2AT}$** | 0.32 | -0.02 | 0.14 | -0.18 | 0.03 | -0.16 | -0.07 | -0.23 |
| **$VO_{2peak}$** | 0.48* | 0.16 | 0.36 | 0.2 | 0.07 | -0.32 | -0.06 | -0.1 |
| **R5** | -0.06 | -0.03 | -0.25 | -0.09 | -0.01 | -0.05 | -0.29 | 0.11 |
| **X5** | 0.12 | -0.06 | 0.22 | -0.05 | -0.01 | -0.07 | 0.16 | -0.23 |
| **R5-19** | 0.02 | -0.14 | -0.27 | -0.09 | -0.17 | -0.11 | -0.23 | 0.05 |
| **AX** | -0.08 | -0.17 | -0.37* | -0.02 | -0.04 | -0.05 | -0.27 | 0.23 |
| **IV antibiotics (total days)** | -0.41** | -0.10 | -0.31* | 0.52*** | 0.43** | 0.27 | 0.19 | 0.38** |
| **Area Under Curve** | | | | | | | | |
| **Pancreatic Insufficiency** | 0.63 | 0.63 | 0.65 | 0.83** | 0.77** | 0.53 | 0.57 | 0.66 |
| **Azithroymycin use** | 0.71* | 0.58 | 0.61 | 0.80*** | 0.80*** | 0.64 | 0.73** | 0.75** |
| **Dornase alfa use** | 0.73* | 0.61 | 0.75* | 0.85** | 0.74* | 0.79* | 0.54 | 0.76* |

Footnote: Spirometry (as GLI z scores): Forced Expiratory Volume in 1 second ($FEV_1$), $FEV_1$/Forced vital capacity ($FEV_1/FVC$), Forced expiratory volume between 25-75% ($FEV_{25-75}$). Multiple breath washout: Lung clearance Index (LCI), $S_{cond}$, $S_{acin}$. CT: Bronchiectasis Index (BEI) and Air Trapping (AT). *$P$<0.05, **$P$<0.01, and ***$P$<0.001.
